# Association between 2019-nCoV transmission and N95 respirator use

**DOI:** 10.1101/2020.02.18.20021881

**Authors:** Xinghuan Wang, Zhenyu Pan, Zhenshun Cheng

**Author notes:** **Corresponding author:** Dr. Xinghuan Wang.

## Abstract

2019-nCoV had caused pneumonia outbreak in Wuhan. Existing evidence have confirmed the human-to-human transmission of 2019-nCoV. We retrospectively collected infection data from 2 January to 22 January at six departments from Zhongnan Hospital of Wuhan University. In our study, we found N95 respirators, disinfection and hand washing can help to reduce the risk of 2019-nCoV infection in medical staffs. Our results call for re-emphasizing strict occupational protection code in battling this novel contagious disease. The risk of 2019-nCoV infection was higher in the open area than in the quarantined area. N95 may be more effective for 2019-nCoV infections.

---

Cases of a novel type of contagious pneumonia were first discovered a month ago in Wuhan. The Centers for Disease Control and Prevention (CDC) and Chinese health authorities have determined and announced that a novel coronavirus (CoV), denoted as 2019-nCoV, had caused this pneumonia outbreak.^1,2^ Existing evidence have confirmed the human-to-human transmission of 2019-nCoV.^3^

We retrospectively collected infection data from 2 January to 22 January at six departments (Respiratory, ICU, Infectious Disease, Hepatobiliary Pancreatic Surgery, Trauma and Microsurgery, and Urology) from Zhongnan Hospital of Wuhan University. Medical staffs in these department follow differential routines of occupational protection: the medical staff in departments of Respiratory, ICU, and Infectious Disease wore N95 respirators, disinfected and clean hands frequently (N95 group); due to people were not enough for the knowledge of the 2019-nCoV in these early days of the pneumonia outbreak, the medical staff in the other three departments wore no medical masks, disinfected and clean hands occasionally (no-mask group). Suspicious cases of 2019-nCoV infection was diagnosed with chest CT and confirmed with molecular diagnosis. In total, 28/58 ([confirmed/suspicious]) 2019-nCoV patients have been diagnosed during the collection period. Patient exposure was significantly higher for the N95 group compared to no-mask group (Table 1) (For confirmed patients: difference: 733%; exposure odds ratio: 8.33)

**Table 1.**
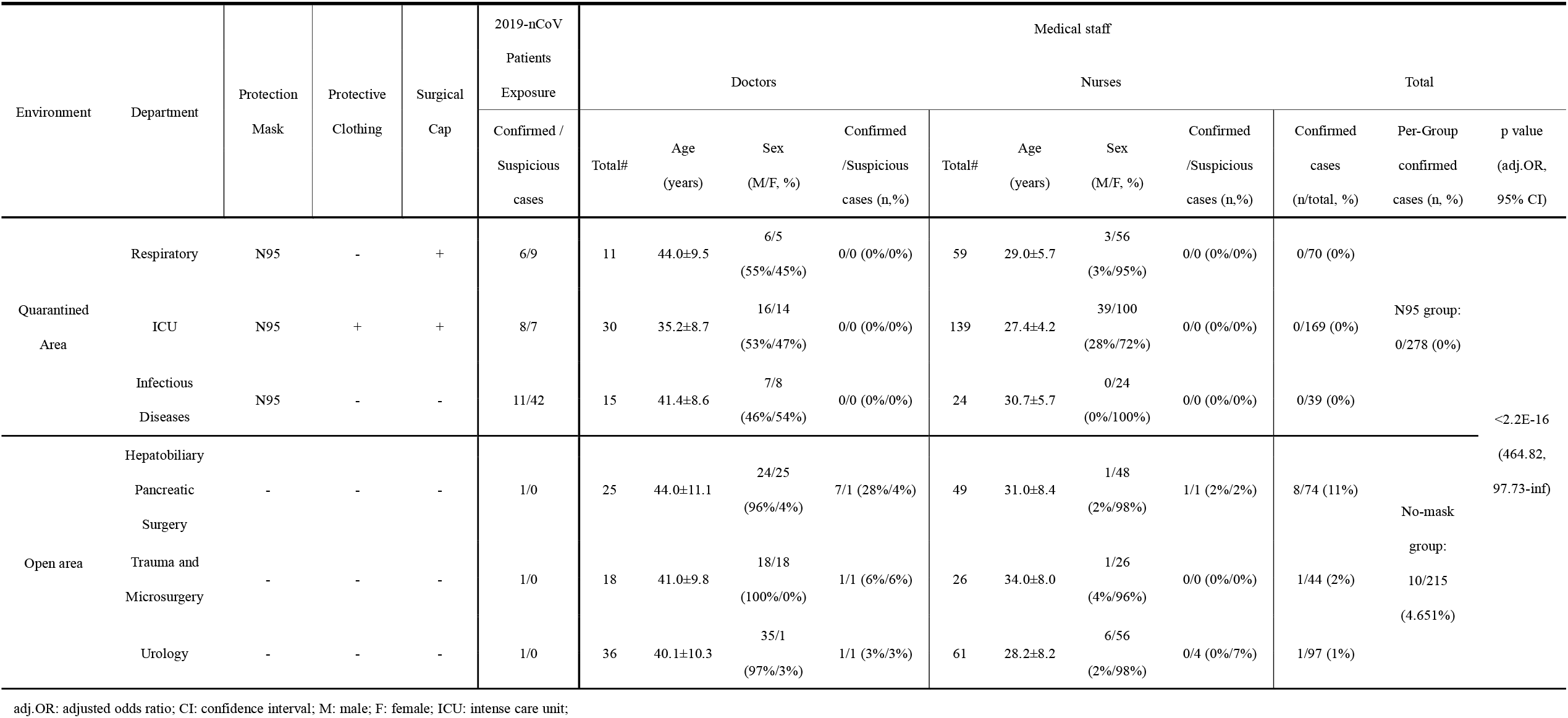
The infection data of patients and medical staff in Zhongnan Hospital of Wuhan University (January 2 to January 22, 2020)

Among 493 medical staffs, zero out of 278 (([doctors+nurse] 56+222) from the N95 group were infected by 2019-nCoV. In stark contrast, 10 out of 213 (77+136) from the no-mask group were confirmed infected (Table 1). Regardless of significantly lowered exposure, the 2019-nCoV infection rate for medical staff was significantly increased in the no-mask group compared to the N95 respirator group (difference: 4.65%, [95% CI: 1.75%-infinite]; P<2.2e-16) (adjusted odds ratio (OR): 464.82, [95% CI: 97.73-infinite]; P<2.2e-16).

Meanwhile, we analyzed the medical staff infection data from Huangmei People’ s Hospital (12 confirmed patients) and Qichun People’ s Hospital (11 confirmed patients) and have observed the similar phenomenon. None of medical staff wearing the N95 respirators and follow frequent routines of disinfection with hand washing were infected by 2019-nCoV.

A randomized clinical trial has reported N95 respirators vs medical masks as worn by participants in this trial resulted in no significant difference in the incidence of laboratory confirmed influenza.^4^ In our study, we found N95 respirators, disinfection and hand washing can help to reduce the risk of 2019-nCoV infection in medical staffs. Interestingly, department with a high proportion of male doctors seem to have a higher risk of infection. Our results call for re-emphasizing strict occupational protection code in battling this novel contagious disease.

The risk of 2019-nCoV infection was higher in the open area than in the quarantined area. N95 may be more effective for 2019-nCoV infections.

We declare no competing interests. This study was supported in part by grants from the Medical Science Advancement Program (Clinical Medicine) of Wuhan University (TFLC2018002).

## Data Availability

N/A

## Reference

1. Hui DS E IA, Madani TA, et al. The continuing 2019-nCoV epidemic threat of novel coronaviruses to global health - The latest 2019 novel coronavirus outbreak in Wuhan, China. Int J Infect Dis 2020;91:264–6.

2. WHO. (2020). Novel Coronavirus (2019-nCoV). https://www.who.int/emergencies/diseases/novel-coronavirus-2019.

3. Xu X, Chen P, Wang J, et al. Evolution of the novel coronavirus from the ongoing Wuhan outbreak and modeling of its spike protein for risk of human transmission. Sci China Life Sci. in press. https://doi.org/10.1007/s11427-020-1637-5

4. Radonovich LJ, Jr., Simberkoff MS, Bessesen MT, et al. N95 Respirators vs Medical Masks for Preventing Influenza Among Health Care Personnel: A Randomized Clinical Trial. JAMA 2019;322:824–33.

